# Anhedonia, Depression, and Symptom Severity in Obsessive-Compulsive Disorder

**DOI:** 10.1101/2025.05.29.25328570

**Authors:** Brian A. Zaboski, Katherine Jones, Elizabeth F. Mattera

**Author notes:** In the past three years, BZ has consulted with Biohaven Pharmaceuticals and received royalties from Oxford University Press; these relationships are not related to the work described here.

## Abstract

**Background:** Anhedonia, the diminished capacity to experience pleasure, is a transdiagnostic symptom increasingly studied in various psychiatric conditions. While recognized in obsessive-compulsive disorder (OCD), its unique relationship with OCD severity, independent of comorbid depression, remains unclear. This study aimed to investigate the prevalence of anhedonia in individuals with OCD, its association with OCD symptom severity, and whether this association persists after controlling for depressive symptoms.

**Methods:** 227 adult participants with a primary or co-primary OCD diagnosis completed self-report measures for anhedonia, OCD severity, and depressive symptoms. Hypothesis-driven and exploratory analyses included descriptive statistics, hierarchical multiple linear regression, and group comparisons.

**Results:** Clinically significant anhedonia was found in 14.5% of the sample, though mean anhedonia levels were generally low (*M* = 0.96, *SD* = 1.79). A significant positive correlation was initially observed between anhedonia and OCD severity (*r* = .148, *p* = .027). However, in a hierarchical regression model, while depressive symptoms significantly predicted OCD severity (*β* = 0.602, *p* < .001), anhedonia did not explain unique variance (ΔR² < .001, *p* = .931) after controlling for depression.

**Conclusions:** General anhedonia is present in a subset of individuals with OCD and shows an initial correlation with OCD severity. However, this relationship appears to be largely accounted for by comorbid depressive symptoms. These findings underscore the critical importance of controlling for depression when researching OCD, as well as targeting depression when delivering evidence-based interventions for OCD.

---

Obsessive-compulsive disorder (OCD) is a chronic and often debilitating mental health condition characterized by the presence of intrusive, unwanted obsessions and repetitive, ritualistic compulsions (American Psychiatric Association, 2022). Affecting approximately 1-2% of the population over a lifetime (Ruscio et al., 2010), OCD frequently leads to significant impairment in social, occupational, and personal functioning (Reid et al., 2021; Zaboski et al., 2019). Understanding the complex interplay of cognitive, affective, and behavioral mechanisms underlying OCD is crucial for refining etiological models and enhancing treatment efficacy (Storch & McKay, 2014; Zaboski, 2020).

One affective dimension of OCD, which has gained increasing attention across many other psychiatric conditions, is anhedonia, or the diminished capacity to experience pleasure (Pizzagalli, 2022). While long recognized as a core symptom of major depressive disorder (MDD) and prominent in psychotic disorders (Barch et al., 2023; Gandhi et al., 2022), evidence increasingly supports anhedonia as a transdiagnostic construct relevant to numerous psychiatric conditions, including anxiety disorders, eating disorders, substance use disorders, and psychosis (Barkus & Badcock, 2019; Conway et al., 2022; Harrison et al., 2014; Lalousis et al., 2024).

Although less studied in OCD compared to other disorders, anhedonia may be a notable, yet neglected, construct of interest. Initial studies reported clinically significant anhedonia in approximately 28% of individuals with OCD, a rate substantially higher than that observed in non-psychiatric controls (Abramovitch et al., 2014). Data from other investigations have aligned with these findings but have evidenced higher variability with 42% of participants experiencing clinically significant anhedonia (Moore, 2021). Furthermore, Abramovitch et al found that higher OCD severity predicted worse anhedonia, even when controlling for comorbid depressive symptoms, a finding supported by subsequent work (Pushkarskaya et al., 2019).

The presence of anhedonia in OCD may be linked to underlying dysfunctions in reward processing circuitry. Neuroscientific studies point to abnormalities in reward-related brain areas in OCD, such as the striatum and orbitofrontal cortex (Berridge & Kringelbach, 2008; Figee et al., 2011), including findings of attenuated reward anticipation activity (Choi et al., 2014; Figee et al., 2011). Conceptually, the potent negative reinforcement cycle maintaining OCD—where compulsive behaviors temporarily reduce obsession-related distress (Craske et al., 2008, 2014)—might overshadow or interfere with naturally rewarding activities, thereby contributing to anhedonia (Abramovitch et al., 2014).

Despite this emerging evidence, several critical aspects of the relationship between anhedonia and OCD remain underexplored. Most importantly, the extent to which anhedonia uniquely contributes to OCD severity independent of the high rates of comorbid depression (Ruscio et al., 2010) requires further validation, particularly as some research has failed to detect this unique association after controlling for depressive symptoms (Moore, 2021). Clarifying this is essential for determining whether anhedonia is an epiphenomenon of co-occurring depression or a distinct feature intertwined with OCD pathophysiology itself.

The present study aims to address these gaps with a well-characterized sample of individuals diagnosed with OCD. Our primary objectives were: (1) To document the prevalence and severity of general anhedonia in our OCD sample; (2) To examine the zero-order association between general anhedonia and overall OCD severity (DOCS total score); and (3) To test whether this association persists after statistically controlling for depressive symptom severity (BDI total score). Based on Abramovitch et al. (2014) and Pushkarskaya et al. (2019), we hypothesized that anhedonia would be prevalent and elevated in this OCD sample, that higher levels of anhedonia would be significantly positively correlated with OCD severity, and that the positive association between anhedonia and OCD severity would remain statistically significant even after controlling for comorbid depression.

As an exploratory step, we investigated relationships between anhedonia and the specific symptom dimensions measured by the DOCS subscales (Contamination, Responsibility for Harm, Unacceptable Thoughts, Symmetry), before and after controlling for depression, to gain initial insights into the specificity of this association within the OCD phenotype. This study seeks to clarify the role of anhedonia in OCD, contributing to a more comprehensive understanding of the disorder’s affective components and informing future treatment and assessment strategies.

## Methods

### Participants

All procedures received Institutional Review Board approval. Participants were recruited nationally and locally through the Yale OCD Research Clinic. Eligible participants were adults aged 18 - 70 years with a primary or co-primary diagnosis of OCD. OCD was confirmed by (1) either the Mini-International Neuropsychiatric Interview (Sheehan et al., 1998) or Diagnostic Interview for Anxiety, Mood, and OCD and Related Neuropsychiatric Disorders (Tolin et al., 2018), (2) validated by a board-certified psychologist or psychiatrist associated with the study, and (3) severity of ≥ 16 on the Yale Brown Obsessive-Compulsive Scale (YBOCS; Goodman et al., 1989).

Exclusion criteria included any significant medical problem known to affect central nervous system functioning (e.g., epilepsy, stroke, poorly controlled thyroid disease, untreated hypertension), unstable medical conditions, or a history of head injury resulting in loss of consciousness for more than 30 minutes. Further psychiatric exclusions were any other primary diagnosis, active suicidality, a history of psychosurgery, or a current manic state. Substance use disorders were exclusionary with the exception of mild cannabis, mild alcohol, or mild tobacco use disorder.

After completing consent, participants completed self-report scales on REDCap (Harris et al., 2009, 2019), a web-based, HIPAA-compliant system for medical data capture. All participants received compensation ($40) for completing the questionnaires.

### Measures

#### Snaith-Hamilton Pleasure Scale

The Snaith-Hamilton Pleasure Scale (SHAPS; Snaith et al., 1995) was used to measure the prevalence of anhedonia in the sample. This 14-item self-report questionnaire asks participants to reflect on how they have felt in the last few days using a 4-point Likert scale (“Strongly Agree” to “Strongly Disagree”). Items include statements such as, “I would enjoy being with my family or close friends,” and “I would be able to enjoy a landscape or a beautiful view.” The SHAPS uses a dichotomous scoring strategy; each item ranked “disagree” is coded as 1 point, and each item ranked “agree” is coded as 0; possible scores range from 0-14, participants scoring 3 or greater classified as experiencing clinically-significant anhedonia (Snaith et al., 1995). The SHAPS has demonstrated high internal consistency (*ɑ* = 0.82; Nakonezny et al., 2010) as well as acceptable test-retest reliability (*r* = .70; Franken et al., 2007).

#### Beck Depression Inventory-II

The Beck Depression Inventory (BDI-II; Beck et al., 1996) is a 21-item self-report questionnaire used to measure depression. For each item, participants respond to a series of 4-point Likert scale items based on how they felt over the past two weeks. The BDI-II is scored from 0 to 63. Proposed cut-offs are as follows: 0 – 13 (minimal or no depression); 14 – 19 (mild depression); 20 – 28 (moderate depression); and 29 – 63 (severe depression) (Beck et al., 1996). The BDI-II has been extensively validated in a range of populations and psychiatric settings and demonstrates high internal consistency (*ɑ* = 0.91; Dozois et al., 1998) and test-retest reliability (*r* = 0.73 - 0.96; Wang & Gorenstein, 2013).

Following the procedures of Joiner et al. (2003), two subscales were created from the BDI-II. An Anhedonic subscale (aBDI) was derived from items assessing loss of pleasure (item #4), loss of interest (item #12), and loss of interest in sex (item #21). The remaining 18 items, which capture other symptoms of depression, were summed to create a General Depression subscale (gBDI). The gBDI scale was used in the present study. This two-factor approach, distinguishing anhedonic from non-anhedonic depressive symptoms, has been shown in prior studies to have strong convergent and divergent validity even when controlling for the remaining items on the BDI-II (Cogan et al., 2024)

#### Quality of Life Enjoyment and Satisfaction Questionnaire Short Form

The Quality of Life Enjoyment and Satisfaction Questionnaire Short Form (Q-LES-Q-SF) is a 16-item self-report questionnaire developed from the longer 93-item form (Endicott et al., 1993). The Q-LES-Q-SF asks participants to indicate how they felt about a series of life-functioning outcomes—including mood, relationships, economic status, and health—over the past week. Items are scored on a 5-point Likert scale (“very poor” to “very fair”), with total scores ranging from 14 – 70. Higher scores indicate better quality of life. The Q-LES-Q-SF demonstrates high internal consistency (*ɑ* = 0.9) and strong test-retest reliability (*r* = 0.93; Stevanovic, 2011).

#### Dimensional Obsessive-Compulsive Scale

The Dimensional Obsessive-Compulsive Scale (DOCS; Abramowitz et al., 2010) is a 20-item self-report questionnaire with 5-point Likert scales across four core OCD symptom dimensions. These include responsibility for harm, contamination, unacceptable thoughts, and symmetry/completeness. Total scores range from 0 – 80, with > 21 proposed as a cutoff for distinguishing OCD from non-clinical populations (Abramowitz et al., 2010). The DOCs displays high internal consistency (ɑ = 0.9) and moderate test-retest reliability (*r* = 0.66) (Abramowitz et al., 2010). It has good convergent validity with other measures of OCD severity, such as the YBOCS (*r* = 0.54) and the Obsessive-Compulsive Inventory (Foa et al., 2002) (*r* = 0.69) (Abramowitz et al., 2010; Rapp et al., 2016).

### Analytic Plan

#### Preprocessing

Data were compiled from self-report measures administered via REDCap. Individual SHAPS items, originally rated on a 4-point Likert scale (1 = Strongly Disagree to 4 = Strongly Agree), were dichotomously scored consistent with the original validation: responses indicating disagreement with experiencing pleasure (raw scores of 1 or 2) were coded as ‘1’ (anhedonic response), and responses indicating agreement (raw scores of 3 or 4) were coded as ‘0’ (hedonic response). These dichotomized scores were summed to create a total score ranging from 0 to 14, where higher scores indicated greater anhedonia. Following Snaith et al., (1995), participants scoring 3 or greater were classified as experiencing clinically significant anhedonia. Summed scores were calculated on the BDI-II, gBDI, DOCS (total and subscales), and QLES-Q-SF. The gBDI (described above) was utilized for analyses requiring a depression control. Missing data for entire scales were handled by listwise deletion on an analysis-by-analysis basis.

### Statistical Analysis

All statistical analyses were conducted using Python (v3.11.8; Van Rossum & Drake, 2009) with the pandas (McKinney, 2010) library for data manipulation, scipy.stats (Virtanen et al., 2020) for correlations and t-tests/Mann-Whitney U tests, statsmodels (Seabold & Perktold, 2010) for linear regression, and pingouin (Vallat, 2018) for Cronbach’s alpha and partial correlations. Data visualizations were generated using matplotlib (Hunter, 2007) and seaborn (Waskom, 2021). An alpha level of .05 was used for all tests of statistical significance.

Descriptive statistics, including means, standard deviations (SD), and ranges, were calculated for participant age and key clinical measures. Frequencies and percentages were computed for categorical demographic variables. Internal consistency was assessed using Cronbach’s alpha.

To address anhedonia prevalence and its association with OCD severity, prevalence was determined by calculating the percentage of participants meeting the SHAPS criteria for anhedonia (total score ≥ 3). The association between anhedonia and OCD severity was assessed with Pearson correlations. To examine the association between anhedonia and OCD severity while controlling for depression, a hierarchical multiple linear regression was conducted. The DOCS total score served as the dependent variable. gBDI total score (BDI-II total without the three anhedonia items) was entered in Step 1 of the model, and the SHAPS total score was entered into step 2. The significance of the change in R-squared (ΔR²) and the beta coefficient (β) for the SHAPS total score in Step 2 were examined to determine the unique contribution of anhedonia.

Lastly, we conducted exploratory analyses to further understand the relationship between anhedonia and OCD. Firstly, to investigate whether anhedonia relates differently to specific OCD symptom dimensions independent of depression, partial correlations were computed between the SHAPS total and each of the four DOCS subscale scores, controlling for the gBDI. Secondly, the utility of the QLES-Q-SF was explored by examining its raw correlation with SHAPS total and its partial correlation with DOCS total, controlling for gBDI. Finally, to explore differences based on anhedonia status, participants were categorized into “anhedonic” (SHAPS total score ≥ 3) and “non-anhedonic” (SHAPS total score < 3) groups. Independent samples t-tests or Mann-Whitney U tests (where assumptions for t-tests, assessed via Shapiro-Wilk tests for normality and Levene’s test for equality of variances, were not met) were used to compare these groups on DOCS total and subscale scores.

## Results

### Demographics and Descriptive Statistics

The final sample included 227 participants. The mean of participants reporting age (*n* = 197) was 30.47 years (*SD* = 11.20). Of those who reported sex assigned at birth (*n* = 209), the majority were female (69.4%), with 27.8% male, 2.4% preferring not to say, and 0.5% identifying as non-binary/other. Out of 201 participants, most were single (63.2%), followed by married (19.4%), and other domestic partners (10.0%). The sample was predominantly White/Caucasian (77.5% of 209 participants), with 7.7% identifying as Asian, 7.2% as Mixed/More than One Race, and 4.3% as Black/African American. The majority of participants identified as Non-Hispanic (89.0%).

Descriptive statistics for the primary clinical measures are presented in Table 2. The mean SHAPS total score was 0.96 (*SD* = 1.79), suggesting generally low levels of anhedonia in this sample (see Figure 1). Based on the established cutoff of ≥ 3 (Snaith et al., 1995), 14.5% (*n* = 33) of participants were classified as experiencing clinically significant anhedonia (Figure 1).

**Figure 1:**
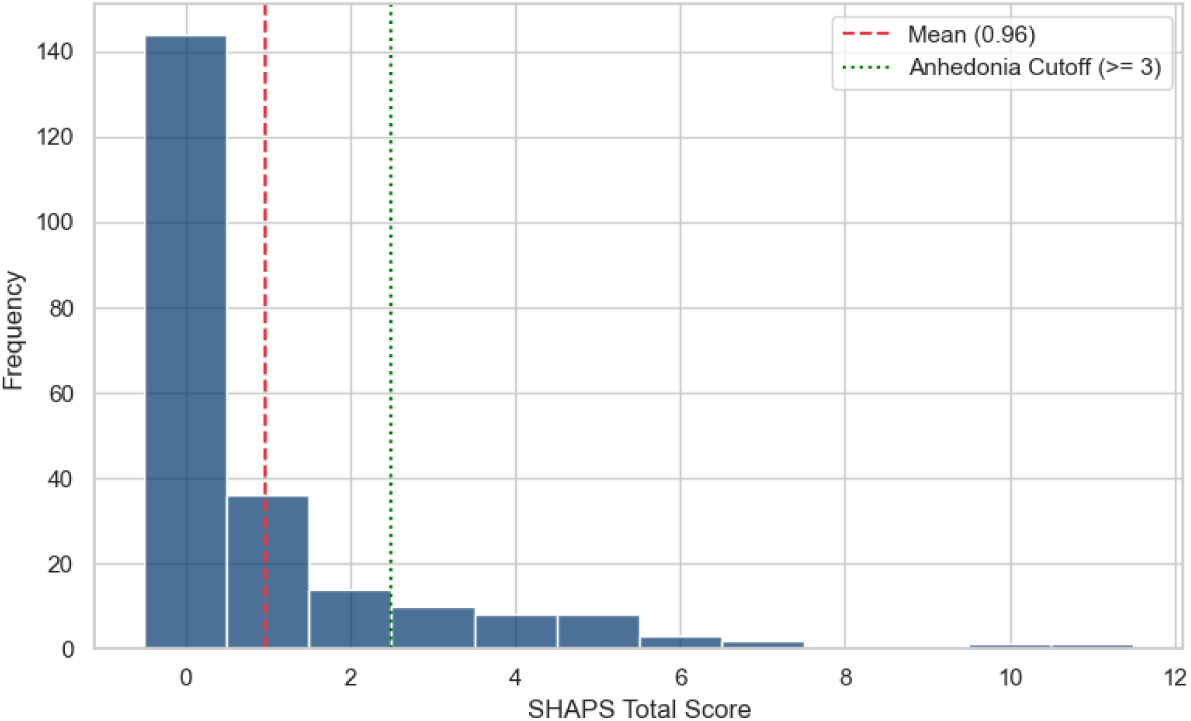
Snaith-Hamilton Pleasure Scale Distribution.

**Table 1:**
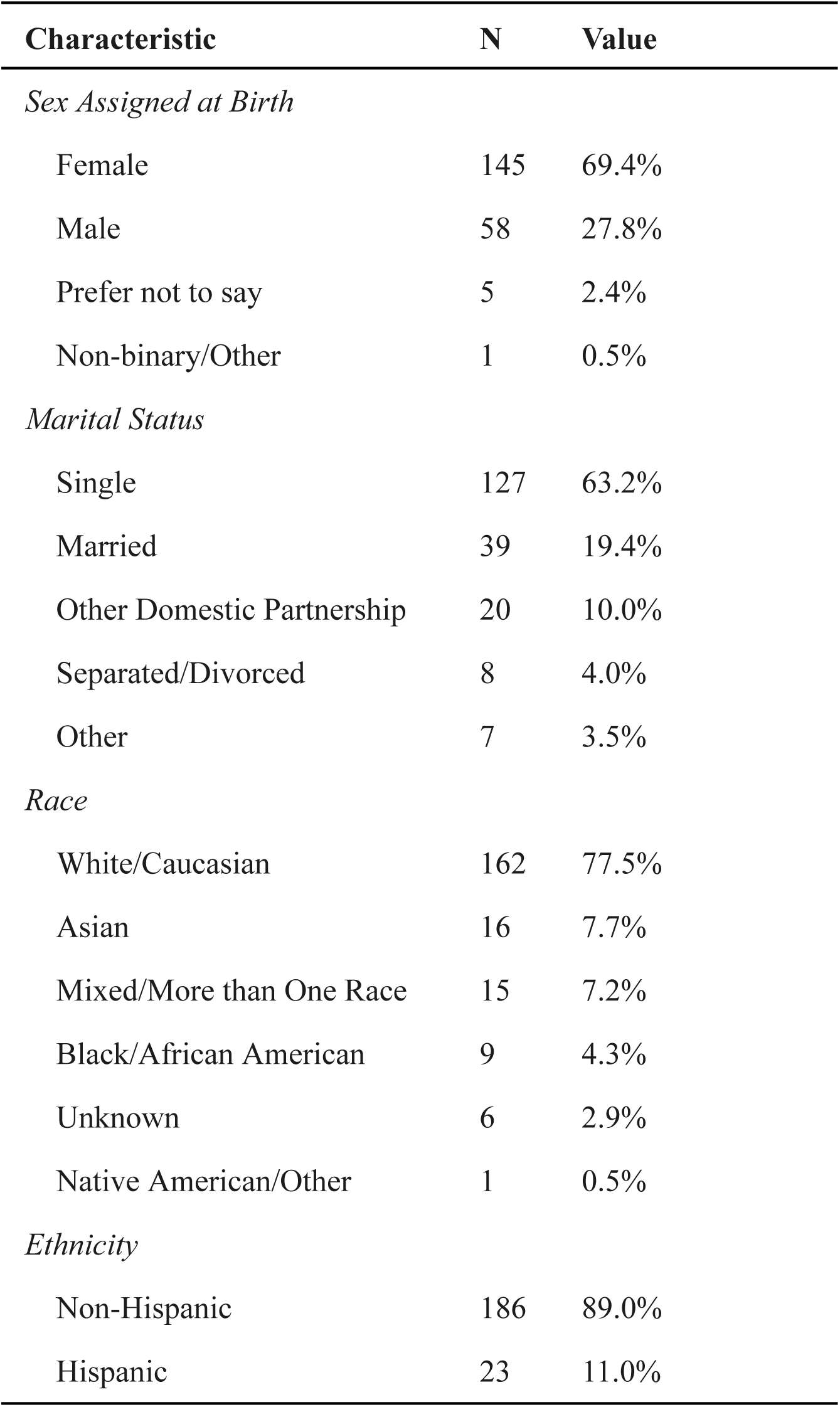
Sample Demographics.

| Characteristic | N | Value |
| --- | --- | --- |
| *Sex Assigned at Birth* |  |  |
| Female | 145 | 69.4% |
| Male | 58 | 27.8% |
| Prefer not to say | 5 | 2.4% |
| Non-binary/Other | 1 | 0.5% |
| *Marital Status* |  |  |
| Single | 127 | 63.2% |
| Married | 39 | 19.4% |
| Other Domestic Partnership | 20 | 10.0% |
| Separated/Divorced | 8 | 4.0% |
| Other | 7 | 3.5% |
| *Race* |  |  |
| White/Caucasian | 162 | 77.5% |
| Asian | 16 | 7.7% |
| Mixed/More than One Race | 15 | 7.2% |
| Black/African American | 9 | 4.3% |
| Unknown | 6 | 2.9% |
| Native American/Other | 1 | 0.5% |
| *Ethnicity* |  |  |
| Non-Hispanic | 186 | 89.0% |
| Hispanic | 23 | 11.0% |

**Table 2:**
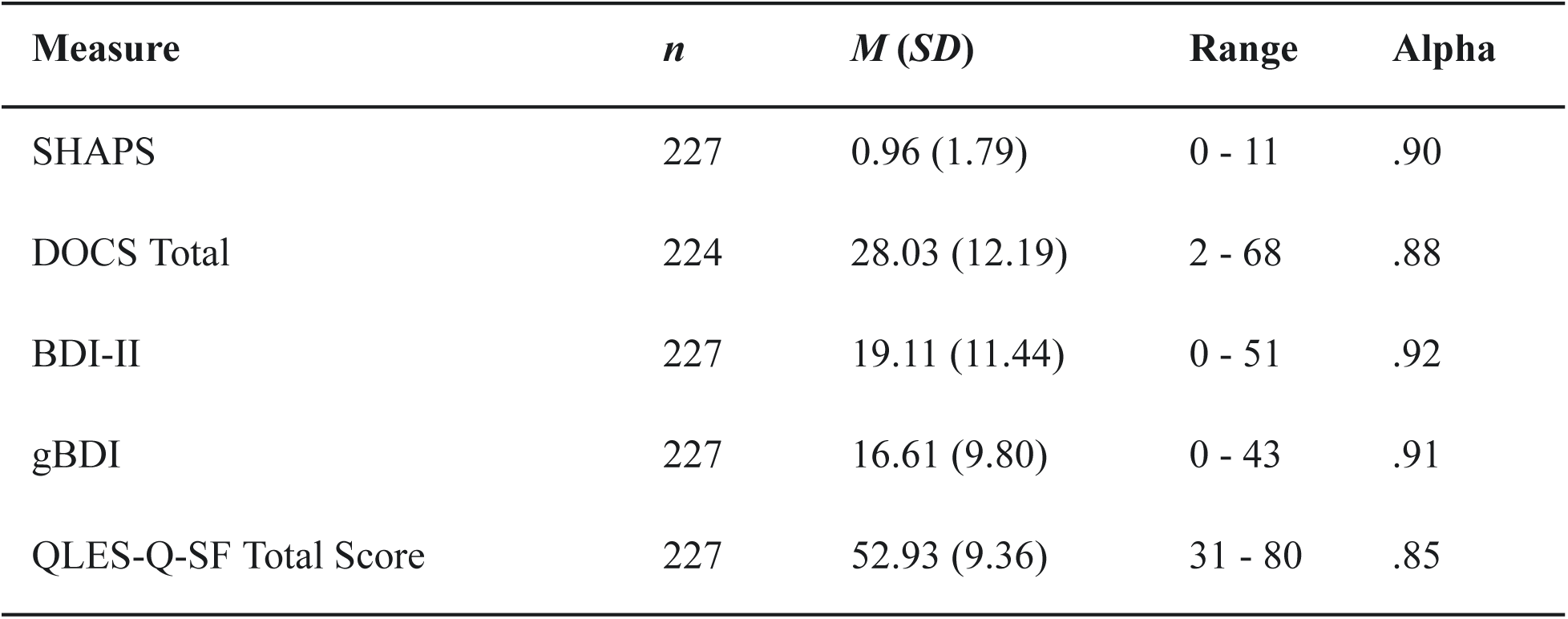
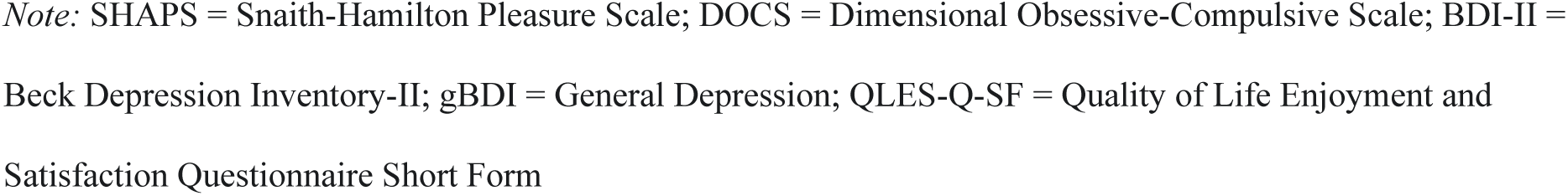
Descriptives Statistic and Internal Consistency.

| Measure | *n* | *M* (*SD*) | Range | Alpha |
| --- | --- | --- | --- | --- |
| SHAPS | 227 | 0.96 (1.79) | 0 - 11 | .90 |
| DOCS Total | 224 | 28.03 (12.19) | 2 - 68 | .88 |
| BDI-II | 227 | 19.11 (11.44) | 0 - 51 | .92 |
| gBDI | 227 | 16.61 (9.80) | 0 - 43 | .91 |
| QLES-Q-SF Total Score | 227 | 52.93 (9.36) | 31 - 80 | .85 |

Internal consistency was also assessed for all primary scales. The SHAPS (based on its 0-3 Likert-style item responses) demonstrated excellent internal consistency (α = .90). Similarly, excellent internal consistency was observed for the BDI-II (α = .92), the DOCS total scale (α = .88), the gBDI (α = .91). The QLES-Q-SF Total Score showed lower, but still very good internal consistency (α = .85). Descriptive statistics and for the clinical measures are contained in Table 2.

### Statistical Analysis

#### Hypothesis Testing

Results on the relationship between anhedonia and OCD severity revealed a significant positive correlation *r* (222) = .148, *p* = .027, indicating that higher levels of anhedonia were associated with greater OCD symptom severity. Bivariate correlations were also examined between anhedonia and the DOCS symptom dimensions (Figure 2). Anhedonia was significantly positively correlated with the Contamination subscale *r* (222) = .137, *p* = .041 and the Unacceptable Thoughts subscale *r* (222) = .166, *p* = .013. Correlations with the Responsibility/Harm subscale *r* (222) = .049, *p* = .470 and the Symmetry subscale *r* (222) = .012, *p* = .856 were not significant.

**Figure 2:**
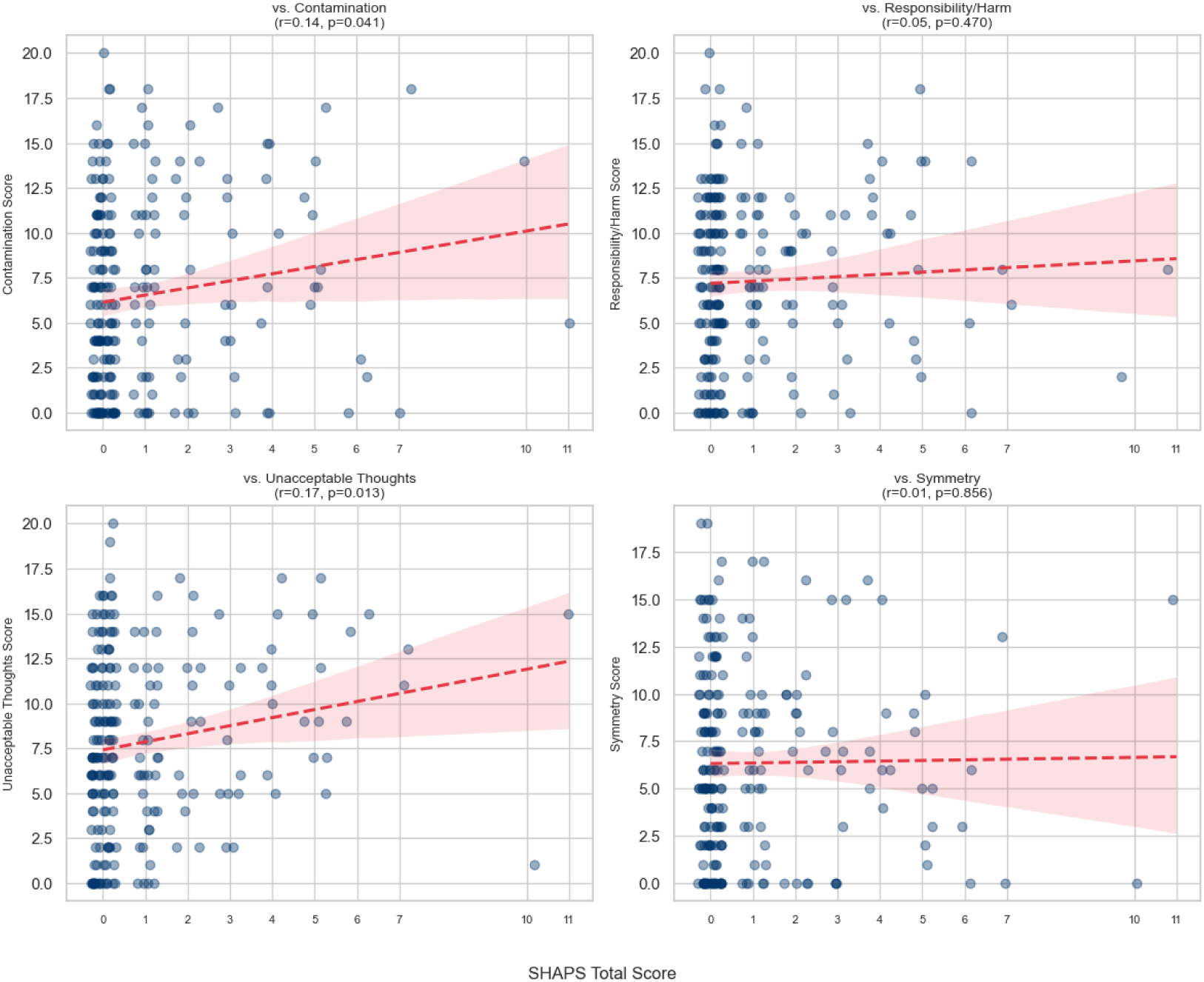
Snaith-Hamilton Pleasure Scale Relationships by Dimensional OCD Scale Dimension.

A hierarchical multiple linear regression was performed to assess whether anhedonia predicted OCD severity beyond general depression severity. In Step 1, gBDI was a significant predictor of the DOCS total score, accounting for 23.7% of the variance *F* (1, 222) = 68.96, *p* < .001; *β* = 0.604, *p* < .001. After adding the SHAPS total in Step 2, the model did not explain significantly more variance in DOCS total score: Δ*R*² = .001, Δ*F*(1, 221) = 0.008, *p* = .931. In the final model, gBDI remained a significant predictor (β = 0.602, *p* < .001), while the SHAPS was not: β = 0.036, *t*(221) = 0.087, *p* = .931.

#### Exploratory Analysis

Partial correlations controlling for gBDI total score were computed to examine the unique relationship between anhedonia and each DOCS subscale. None of these partial correlations were statistically or clinically significant: Contamination *r* (221) = .051, *p* = .445, Responsibility for Harm *r* (221) = - 0.95, p = .158 Unacceptable Thoughts *r* (221) = .046, *p* = .490, and Symmetry *r* (221) = - .012, p = .856.

Additional analyses were conducted with the QLES. The total score demonstrated a significant negative correlation with the SHAPS: *r* (225) = - .355, *p* < .001), indicating that greater life satisfaction and enjoyment was associated with lower levels of anhedonia (see Figure 3 for the raw correlation). The partial correlation between QLES total score and DOCS total score controlling for gBDI was not significant: *r* (221) = - .058, *p* = .389.

**Figure 3:**
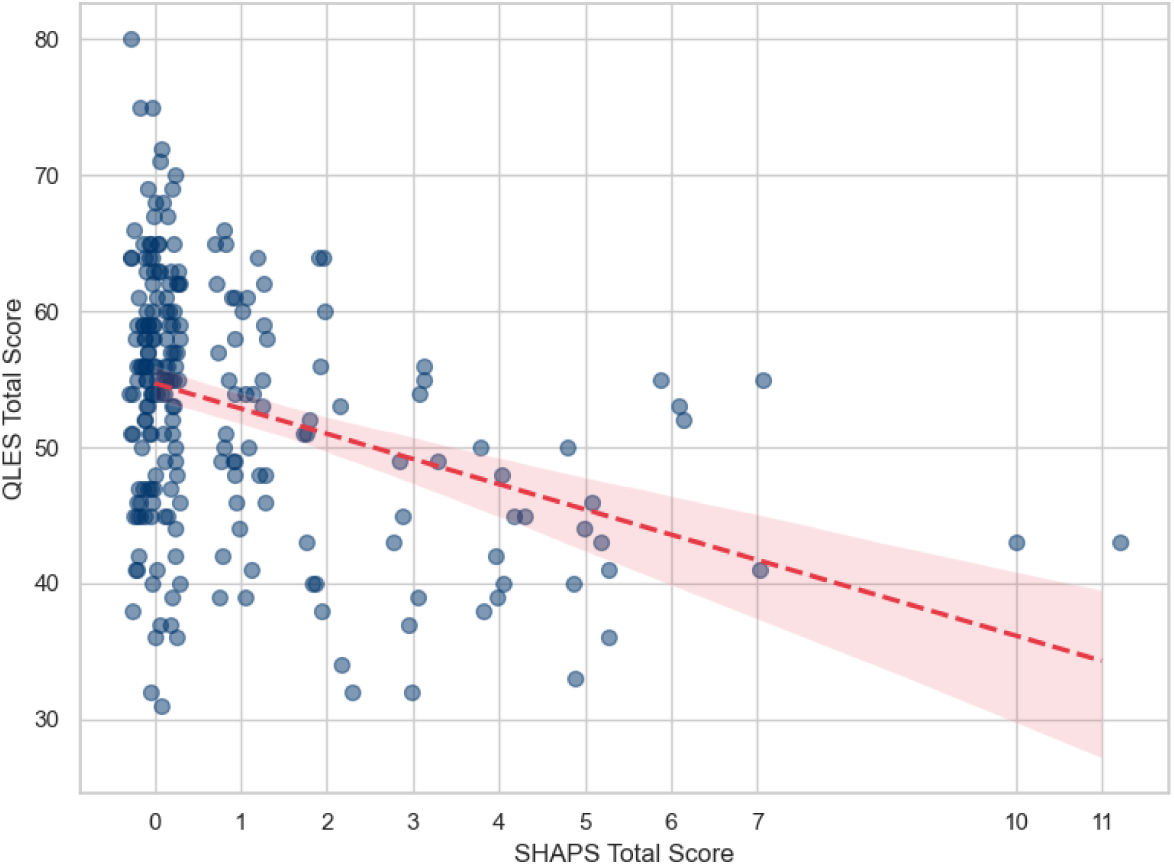
Relationship between the Snaith-Hamilton Pleasure Scale (SHAPS) and Quality of Life Enjoyment and Satisfaction Questionnaire Short Form.

Group comparisons between participants classified as anhedonic (SHAPS total ≥ 3, *n* = 33) and non-anhedonic (SHAPS total score < 3, *n* = 194) revealed significant differences. The anhedonic group reported significantly higher DOCS total scores (*M* = 32.03, *SD* = 10.97) compared to the non-anhedonic group (*M* = 27.34, *SD* = 12.29, *U* = 3922.0, *p* = .025) (Figure 4, left). Specifically, the anhedonic group scored significantly higher on the DOCS Unacceptable Thoughts subscale (*M* = 9.67, *SD* = 4.54) than the non-anhedonic group (*M* = 7.53, *SD* = 4.88), *U* = 3937.0, *p* = .022 (Figure 4, right). No significant group differences were found for the DOCS Contamination (*p*=.114), Responsibility for Harm (*p*=.336), or Symmetry (*p*=.920) subscales.

**Figure 4:**
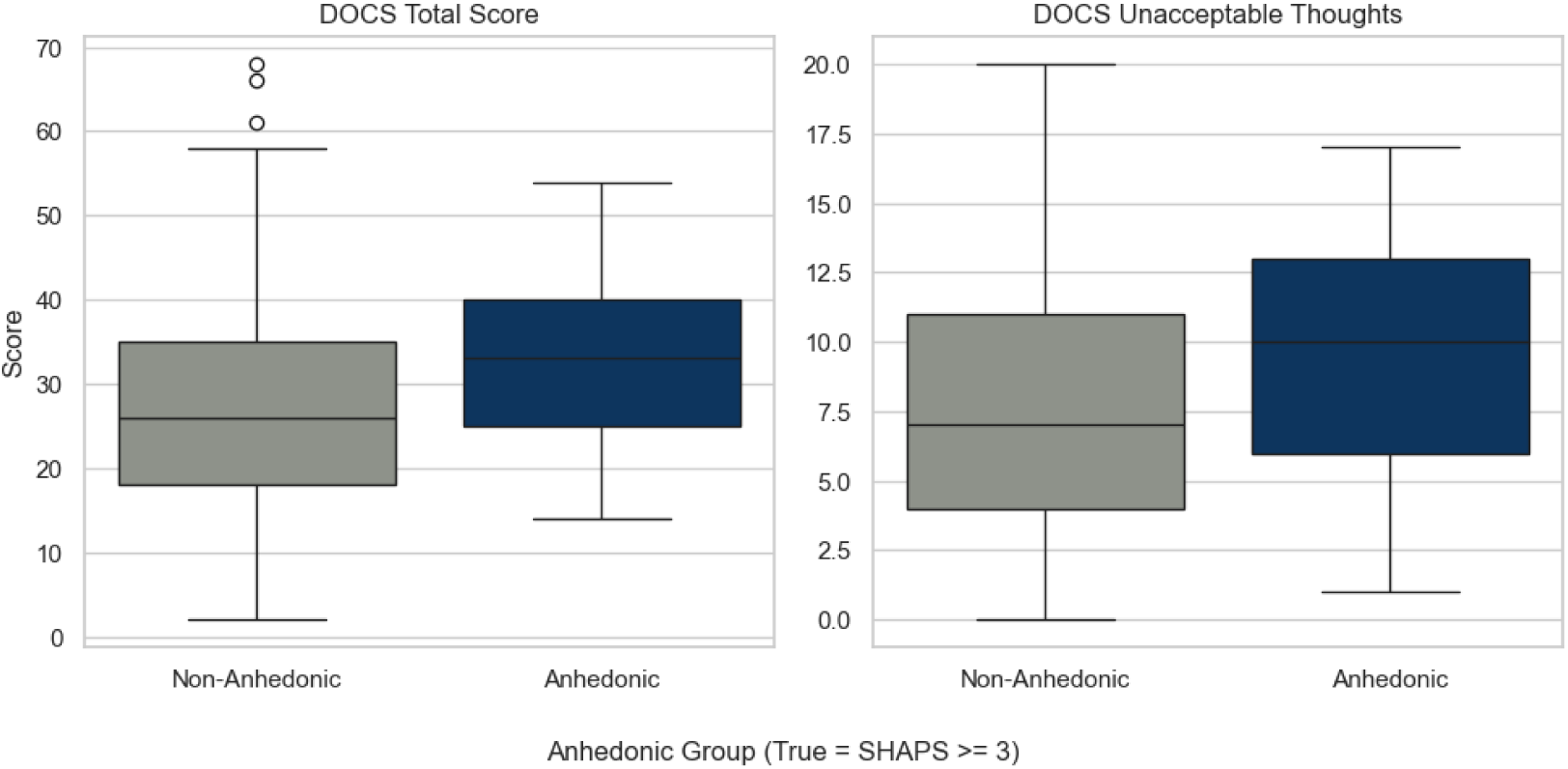
Group Comparisons for Anhedonic and Non-Anhedonic Individuals on the Snaith-Hamilton Pleasure Scale (SHAPS) and Dimensional OCD Scale.

To further investigate whether these group differences were independent of depression, multiple regression analyses were conducted, predicting DOCS Total and DOCS Unacceptable Thoughts scores from the Anhedonia Group status while controlling for gBDI. For DOCS Total score, after controlling for BDI-II (which remained a significant predictor, β = 0.597, *p* < .001), the Anhedonia Group status was no longer a significant predictor (β = 0.709, *p* = .733).

Similarly, for DOCS Unacceptable Thoughts score, after controlling for BDI-II (β = 0.205, *p* < .001), the Anhedonia Group status was not significant (β = 0.767, *p* = .373). These results indicate that the observed differences in overall OCD severity and Unacceptable Thoughts between the anhedonic and non-anhedonic groups were accounted for by differences in comorbid general depression symptoms.

## Discussion

This study investigated the prevalence of anhedonia and its unique association with symptom severity in a well-characterized sample of individuals with OCD. Further, we explored the relationship between anhedonia and specific OCD symptom dimensions. Consistent with previous research indicating anhedonia as a relevant symptom in OCD (Abramovitch et al., 2014; Moore, 2021; Pushkarskaya et al., 2019), we found that 14.5% of the current sample met criteria for clinically significant anhedonia. However, the overall mean SHAPS score suggested generally low levels of anhedonia across the sample. Initial zero-order correlations supported our hypothesis that higher general anhedonia would be associated with greater overall OCD severity. Exploratory analyses also revealed zero-order correlations between anhedonia and the Contamination and Unacceptable Thoughts dimensions of OCD.

Unexpectedly, the positive association between anhedonia and OCD severity was small and non-significant after controlling for comorbid depression. The hierarchical multiple linear regression demonstrated that while depressive symptoms significantly predicted OCD severity, general anhedonia did not add unique predictive value. This finding was further corroborated by partial correlation analyses, which showed no significant unique associations between anhedonia and any of the four DOCS subscale scores after controlling for general depression.

These results contrast with earlier findings that anhedonia predicted OCD severity even after accounting for depression (Abramovitch et al., 2014; Pushkarskaya et al., 2019). However, our findings align with other preliminary work suggesting the link might be fully explained by depression (Moore, 2021). The additional regression results further clarified this picture: While simple group comparisons showed that participants classified as anhedonic reported higher overall OCD severity and higher Unacceptable Thoughts scores, these differences became non-significant after controlling for depression. This strongly suggests that the observed differences in OCD severity between anhedonic and non-anhedonic individuals in this sample were primarily driven by comorbid depressive symptoms.

Several factors might explain the discrepancy between our findings and Abramovitch et al. (2014) and Pushkarskaya et al. (2019). First, sample characteristics differ. For instance, Abramovitch et al. (2014) reported a considerably higher rate of comorbid depression (60.2%) compared to our sample (though our study did not collect full diagnostic data for comparison, descriptive BDI-II scores were close to the moderate range). It is plausible that anhedonia only exerts a unique influence on OCD symptoms when comorbid depression reaches a certain diagnostic threshold or level of severity. Second, differences in measurement could play a role. While both our study and Abramovitch et al. used the SHAPS, Pushkarskaya et al. used a BDI-derived anhedonia subscale with three items. Furthermore, in this study OCD severity was assessed using the DOCS, whereas prior studies often used the Y-BOCS. While all are valid approaches, they capture symptom severity differently.

Additionally, the specific facet of anhedonia measured might be critical. The SHAPS primarily taps general and consummatory aspects of pleasure (Snaith et al., 1995). Research in OCD and related disorders suggests potential dissociations between consummatory and anticipatory pleasure (Li et al., 2019; Sherdell et al., 2012), as well as between physical and social anhedonia (Chapman et al., 1980; Moore, 2021; Xia et al., 2019). Studies using the Temporal Experience of Pleasure Scale (TEPS; Gard et al., 2006) found OCD patients exhibited deficits in consummatory but not anticipatory pleasure, distinct from patients with major depressive disorder (MDD) who showed deficits in both (Li et al., 2019). Other work suggests links between social anhedonia and OCD (Moore, 2021), and specific brain activity alterations related to anhedonia severity in OCD, particularly involving the superior temporal gyrus (associated with social perception) and medial prefrontal cortex (Xia et al., 2019). It is conceivable that the SHAPS, by focusing on general/consummatory pleasure, may not capture an aspect of reward dysfunction more uniquely relevant to OCD pathophysiology, such as deficits in anticipatory pleasure or social reward processing. Our exploratory analysis using the QLES-Q-SF yielded a negative correlation with anhedonia, but no unique association with OCD severity after controlling for depression. These parallel our SHAPS findings and prior analyses showing weak relationships between quality of life, OCD, and anxiety severity after controlling for depression (Molinari et al., 2019; Zaboski et al., 2019). This might suggest that broader measures of pleasure/enjoyment are strongly tied to mood state but less specifically to core OCD mechanisms independent of that mood state.

The conceptualization of OCD, particularly regarding reward processing, also warrants consideration. While often viewed through an anxiety/threat lens, models proposing OCD as a behavioral addiction emphasize dysfunctional reward processing, potentially involving negative reinforcement (relief from anxiety) overriding natural rewards (Abramovitch et al., 2014; Choi et al., 2014; Figee et al., 2011). Neuroimaging studies point to altered activity in reward-related regions like the nucleus accumbens and orbitofrontal cortex during reward anticipation in OCD (Choi et al., 2014; Figee et al., 2011). Some studies suggest impaired associative learning from external feedback in OCD (Nielen et al., 2009), potentially hindering the ability to update behavior based on rewarding outcomes. Anhedonia could theoretically arise from or contribute to this cycle. However, our finding that general anhedonia does not uniquely predict OCD severity complicates this picture, suggesting that if reward dysfunction is core to OCD independent of depression, it might manifest in ways not fully captured by the SHAPS, perhaps relating more to anticipatory processes, specific learning deficits, or the processing of relief as a reward. The finding by Grassi et al., (2020) that anhedonia scores in OCD were related to depression/anxiety symptoms but not in patients with gambling disorder, aligns with our results suggesting anhedonia in OCD is tightly coupled with comorbid affective symptoms.

### Clinical Implications

While anhedonia is present in a subset of individuals with OCD, its severity may not be a direct target for reducing OCD symptoms *themselves*, unless comorbid depression is also a primary treatment focus (see Storch & McKay, 2014). Interventions aimed at increasing pleasure and engagement might improve overall quality of life and potentially alleviate depressive symptoms. However, based on these findings, clinicians might not expect a direct corresponding decrease in OCD severity solely from targeting general anhedonia. Accurate assessment of both depressive symptoms and anhedonia remains crucial for comprehensive treatment planning in OCD.

### Limitations and Future Directions

This study has several limitations. Its cross-sectional nature precludes causal inferences. Reliance on self-report measures also carries inherent biases. Future research should incorporate multiple methods, including diverse self-report scales capturing different facets of anhedonia (e.g., TEPS for anticipatory/consummatory, Chapman scales for physical/social; Chapman et al., 1976) as well as clinician-rated and behavioral measures. Using clinician-administered OCD scales like the Y-BOCS alongside self-report like the DOCS would also strengthen findings. The relatively low prevalence of clinical anhedonia (14.5%) despite the significant correlations we found warrants further investigation in samples with higher anhedonia rates. Finally, mechanistic studies using neuroimaging or behavioral reward tasks are essential to directly probe reward processing dysfunction and its specific relationship to different facets of anhedonia and OCD symptoms, independent of depression. Exploring potential variations across OCD symptom dimensions warrants further investigation, although our initial partial correlations did not suggest dimension-specific links independent of depression.

### Conclusion

While general anhedonia is correlated with OCD severity and present in a meaningful minority of individuals with OCD, this association appears to be largely accounted for by comorbid depressive symptoms. These findings highlight the critical importance of controlling for depression when examining affective constructs in OCD and suggest that general anhedonia, as measured by the SHAPS, may not be uniquely associated with the severity of core OCD symptoms. Future research should employ diverse methodologies and measures targeting specific facets of reward processing and anhedonia to further elucidate the complex relationship between affective disturbance and OCD pathophysiology.

## Data Availability

All data produced in the present study are available upon reasonable request to the authors.

